# Evaluation of a Modified Early Warning Score (MEWS) adjusted for COVID-19 patients (CEWS) to identify risk of ICU admission or death

**DOI:** 10.1101/2020.10.22.20217539

**Authors:** Linda Smid, Lano Osman, Sandra M. Arend, Stefanie de Groot, Mark G.J. de Boer

## Abstract

The commonly used Modified Early Warning score (MEWS) may poorly predict deterioration in COVID-19 patients. Therefore, an adjusted MEWS for COVID-19 patients (CEWS) was constructed. CEWS exceeded MEWS at all time points and impending death or intensive care unit (ICU) admission was strongly correlated with a persistently high CEWS.

## Introduction

The clinical spectrum of COVID-19 varies from mild upper respiratory symptoms to acute respiratory distress syndrome and death [1]. Estimates are that 20% of patients infected with SARS-CoV-2 require hospital admission and a critical course has been described in about 15% of these hospitalized patients [2–4]. The validated Modified Early Warning Score (MEWS), which is based on a set of vital parameters, is commonly used to monitor the patient’s condition objectively and is aimed at early detection of clinical deterioration [5, 6]. An increase of the MEWS often results in re-assessment of the patient’s condition and/or consultation of intensive care staff. However, the MEWS contains only few respiratory parameters and during the pandemic surge we observed that the MEWS did not always seem to accurately reflect the deterioration of the respiratory status of COVID-19 patients. For example, on our COVID isolation ward several patients were admitted with a MEWS between 2-4, while at the time of ICU admission or death their MEWS was comparable. We hypothesized that the inclusion of additional respiratory parameters in an adjusted Early Warning Score for COVID-19 patients (CEWS) would show a stronger correlation with mortality or the need for subsequent ICU admission than the MEWS.

## Methods

All adult (≥18 years old) patients consecutively admitted with a proven COVID-19 infection from March 18^th^ to April 30^th^ 2020, were considered eligible for this study. A proven COVID-19 infection was defined as a positive SARS-CoV-2 PCR on a respiratory sample and/or a CT-scan of the thorax reported to fulfill CO-RADS 5 criteria [7]. Exclusion criteria were: direct transfer from another hospital to our hospital’s ICU, availability of less than three complete sets of vital parameters, absence of a complete set of vital parameters <6 hours prior to the endpoint, or discharge with palliative care. Patient groups were established based on clinical course and outcome. Primary endpoints were ICU admission and death, which defined case patients. If these endpoints were not reached, i.e. in control patients, the moment with the highest MEWS score during admission - the clinical nadir - was used as comparative endpoint. We obtained data about age, gender and all vital parameters that were measured during the 48 hours before reaching the endpoint (cases) or comparative endpoint (controls). Consent for use of clinical data was obtained through an opt-out procedure. This study was approved by the Institutional Review Board of the LUMC for observational studies.

The MEWS was incorporated in standard care in our hospital for >2 years and registered in the medical file by the nursing staff. The CEWS consists of an adjusted MEWS by further dividing the oxygen saturation level into categories and adding the amount of oxygen supplementation per minute as a categorical parameter (**Figure 1**). For each time point with a complete set of parameters, the MEWS and CEWS were calculated.

**FIGURE 1.**
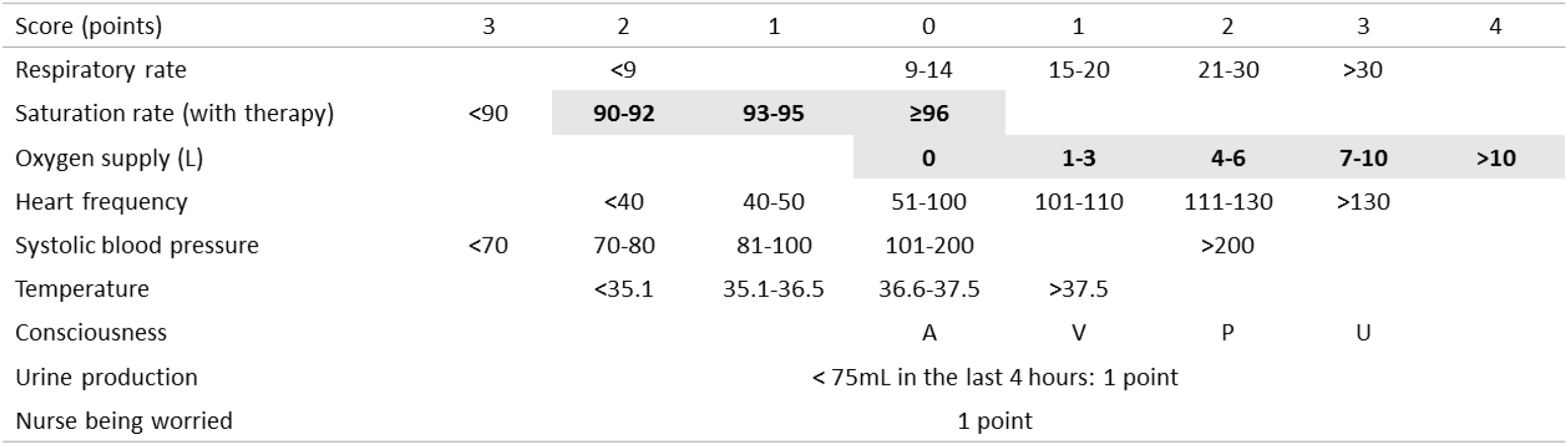
MEWS and CEWS scoring table. A = Alert V = Verbal reaction P = Reaction to pain U = Unresponsive The current MEWS was adjusted, by: 1. further dividing the oxygen saturation level into categories (bold, shaded cells) and 2. adding the amount of oxygen supplementation per minute as parameter, also into categories (bold, shaded cells).

The 48-hour time period of interest was divided in the following time periods: 48-36, 36-24, 24-12, 12-6, 6-3 and 3-0 hours prior to the endpoint or comparative endpoint. The maximum MEWS and CEWS values of the last three available time periods prior to the endpoint were used for the analyses. A repeated measurements ANOVA model was applied to compare values of MEWS and CEWS over time and between case- and control patients. Mauchly’s test was used to assess the sphericity of the data. If indicated, the Greenhouse-Geisser or Huynh-Feldt corrections were applied to interpret the results. The data was analyzed using Statistical Package for Social Sciences (SPSS) software™ 23.0 (IBM Corp., Armonk, NY, USA).

## Results

One hundred-and-twenty-five patients were evaluated for inclusion, of which 84 patients were excluded due to the following reasons: 64 patients were transferred from an ICU elsewhere, 19 patients had less than three complete sets of vital parameters and one case was discharged with palliative care. Hence a total of 22 case- and 19 control patients were included. The mean age and age-range was 73 (52-89) and 66 (26-88) years for cases and controls, respectively. In the case group 13/22 (59%) patients were male, versus 11/19 (58%) in the control group. The diagnosis was primarily based on a positive PCR (N= 38) and on a CT with a CO-RADS 5 in three patients.

The assumptions for sphericity of the data were met for CEWS, but not for MEWS data. For the total study population, the repeated measurements ANOVA analysis showed that within individual patients, maximum values of both MEWS (p=0.045 after Greenhouse-Geisser correction) and CEWS (p=0.026) differed significantly over the three time periods. In the pairwise analyses, this difference was shown to be driven by an increase of the maximum warning score value from the penultimate time period to the last time period before the endpoint (p=0.034 and p=0.022 for MEWS and CEWS, respectively). The numerical increases of MEWS or CEWS were not correlated with attainment of a primary endpoint (death or ICU admission), i.e. with being a case patient. However, for both MEWS and CEWS, during all three time periods the mean scores were consistently higher in cases compared to controls (p=<0.001), with CEWS being the most discriminative (**Figure 2c**).

**FIGURE 2.**
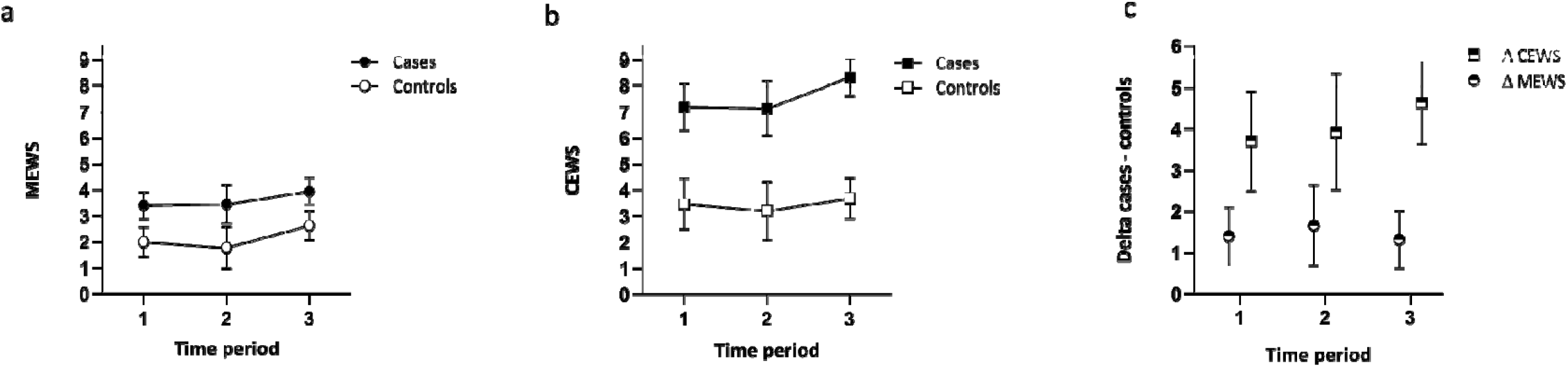
Mean values of MEWS and CEWS over time. Mean maximum values of the MEWS (2a) and CEWS (2b) in cases and controls in the last three time periods prior to the endpoint (95% confidence interval). Figure 2c shows the mean difference (delta) between cases and controls for MEWS and CEWS (95% confidence interval).

## Discussion

We evaluated the time course of MEWS in admitted COVID-19 patients and made a proposal for a new adjusted score for this patient group. We found a significant increase in both MEWS and CEWS during the 48 hours before the endpoint. However, the average increase was similar for the case- and control patients. While the kinetics of MEWS and CEWS thus lacked discriminatory power, the absolute value of both scores was significantly higher in the cases than in controls at all time points, with the difference between CEWS in cases and controls being most striking. Interestingly, during the observed time period we noticed a persistently elevated early warning score in the cases, for both MEWS and CEWS. This strongly suggests that not just the value of MEWS or CEWS, but a persistently increased early warning score (‘the area under the MEWS/CEWS curve’) could predict ICU admission and/or death. This finding is consistent with the clinical observation in COVID-19 that patients can seem to be more or less stable at a high respiratory rate and oxygen requirement for some time, followed by sudden exhaustion with rapid deterioration to respiratory failure [4, 8, 9]. This compensatory respiratory capacity differs between individuals, and most probably depends on pre-existing fitness and pulmonary reserves.

In clinical practice, various early warning scores are used to predict clinical deterioration. In our hospital the MEWS is routinely used. It has been shown that the MEWS can significantly reduce the occurrence of cardiopulmonary arrests and in-hospital mortality and lower the risk of unplanned ICU admissions [6]. Thus far, only a few cohort studies have been conducted investigating the predictive value of an early warning score in patients with a COVID-19 infection. These studies all contained a limited number of critically ill patients, and only calculated the early warning score at the time of admission. Two retrospective studies showed a significantly higher National Early Warning Score (NEWS) and higher NEWS2 (version 2 of the score) at the time of admission in patients who eventually required ICU care [10, 11]. The NEWS2 seemed to better predict a critical course than CURB-65 (score for pneumonia severity) and qSOFA (quick sequential organ failure assessment) [10]. In comparison with the MEWS, NEWS(2) contains a category O2 supply and a more detailed subdivision of the saturation level. Hu et al. compared the MEWS with the Rapid Emergency Medicine Score (REMS) in predicting in-hospital mortality for critically ill COVID-19 patients, where the REMS seemed superior, though critically ill patients had a low mean MEWS of 2 on admission [12]. Finally, Peng et al. prospectively described a cohort of 200 patients of whom 12 eventually needed artificial ventilation or died [13]. Given the limited number of serious events, the NEWS had a good sensitivity and mediocre specificity for predicting deterioration in patients admitted with a COVID-19 infection (0.92 and 0.57, respectively).

A limitation of our study is the modest sample size. In addition, information bias could have played a role since vital parameters are measured more often in seriously ill patients. As a result, a more accurate course of the MEWS and CEWS can be expected in case patients as compared to less ill control patients.

Based on our results, a persistently elevated early warning score, in particular CEWS, indicates a high risk of an upcoming life-threatening event. The difference between the CEWS in cases versus controls exceeded the difference in the MEWS, and seems to be a more reliable warning score of the need for ICU admission or mortality. Further prospective studies are needed to validate the predictive value of a persistently high MEWS or CEWS in patients with a COVID-19 infection.

## Data Availability

All data referred to in the manuscript is available.

## Funding

There are no funding sources.

## Conflict of interest

All authors declare to have no conflicts of interests.

## Notes

### Competing Interest Statement

The authors have declared no competing interest.

### Clinical Trial

Retrospective cohort study.

### Author Declarations

The Covid-19 science committee for non-WMO (Medical Scientific Research with Human Subjects Act) research assessed the investigation and issued a statement of no objection to the conduct of the investigation.

